# An Explainable Host Genetic Severity Predictor Model for COVID-19 Patients

**DOI:** 10.1101/2023.03.06.23286869

**Authors:** Anthony Onoja, Francesco Raimondi, Mirco Nanni

## Abstract

Understanding the COVID-19 severity and why it differs significantly among patients is a thing of concern to the scientific community. The major contribution of this study arises from the use of a voting ensemble host genetic severity predictor (HGSP) model we developed by combining several state-of-the-art machine learning algorithms (decision tree-based models: Random Forest and XGBoost classifiers). These models were trained using a genetic Whole Exome Sequencing (WES) dataset and clinical covariates (age and gender) formulated from a 5-fold stratified cross-validation computational strategy to randomly split the dataset to overcome model instability. Our study validated the HGSP model based on the 18 features (i.e., 16 identified candidate genetic variants and 2 covariates) identified from a prior study. We provided post-hoc model explanations through the ExplainerDashboard - an open-source python library framework, allowing for deeper insight into the prediction results. We applied the Enrichr and OpenTarget genetics bioinformatic interactive tools to associate the genetic variants for plausible biological insights, and domain interpretations such as pathways, ontologies, and disease/drugs. Through an unsupervised clustering of the SHAP feature importance values, we visualized the complex genetic mechanisms. Our findings show that while age and gender mainly influence COVID-19 severity, a specific group of patients experiences severity due to complex genetic interactions.

## 1. Introduction

The severe acute respiratory syndrome coronavirus 2 (SARS-CoV-2) has continued to pose a great threat to humanity ever since its first outbreak in late 2019. The SARS-CoV-2 viral strand causes the new coronavirus of 2019 popularly known as COVID-19 which has claimed millions of lives. The disease is widely characterized by a spectrum of clinical severity, suggesting a complex and highly dynamic host response in patients^1^. Host (human) genetic variation associated with severity susceptibility or infection might provide clues to effective points to develop therapy or even preventive measures to intervene to develop medicine and vaccine against SARS-CoV-2 infection^2^. Most especially, the scientific community is of kin interest that such findings provided by genetic human variations could give important clues where existing drugs may be repurposed for effective therapy against SARS-CoV-2 infection and life-threatening COVID-19 disease^3^. Also, we might be able to spot groups of individuals in the human population that might be at unusually high risk and need to be protected or might have innate protection against the SARS-CoV-2 infection^4–7^. The SARS-CoV-2 genetic severity and susceptibility can also manifest themselves in rare genetic mutations which can cause healthy individuals to have a life-threatening response to COVID-19 disease^6^. Comorbidities such as diabetes, hepatitis, HIV, kidney-related problems, age, and gender have been observed in several clinical studies to have strong ties with patients’ severity and susceptibility to the disease^7,8^. Some hosts are more susceptible to developing a severe disease probably due to modulated influence of genetics, environment, and risk factors.

There is a knowledge gap as to why the response to COVID-19 infection varies so much from patient to patient. The study of human genetics to diseases from several studies has pointed out some links to the severity of the disease among some groups of patients ^9^. For example, in some cases healthy and young patients with no prior existing medical conditions when exposed to the disease developed severe symptoms and some even succumbed to death from the disease. Emerging evidence suggests that asymptomatic patients mount a weaker immune response to the COVID-19 virus^10–12^. There are some complex genetic interactions with the disease on the host side that can help to explain the variability in COVID-19 severity susceptibility and outcomes among patients^13^. Vital information as to why the disease differs greatly between people might lie in their DNA (e.g., variations in immune-related genes). Gene expression identifies patterns within human immune cells and may also play a key role in determining how the host immune system interacts with the virus. Examining genomes of patients who have a severe response to COVID-19 becomes necessary to further understand these complex interactions that are crucial to shed more light on understanding the biology of the disease, selecting drugs for repurposing – and knowing patients who are most at risk or providing some sorts of protection against the infection^14,15^.

Most scholarly works so far utilized the Geno-Wide Association studies (GWS) approach to identify pointer chromosome loci, variants, and genes that linked COVID-19 susceptibility severity in patients^16,17^. These studies have been vital in establishing crucial genetic variants linked to the disease, for example, the study of Q. Zhang *et al*.,^18^ used a candidate gene approach to identify patients with severe COVID-19 who have mutations in genes involved in the regulation of type I and III IFN immunity. Their studies found enrichment of these genes in patients and concluded that genetics may determine the clinical course of the infection. However, the GWS approach usually adopts more stringency measures for filtering genetic variants associated with a disease. For example, the threshold of small p-values is usually lowered to identify highly significant genetic variants with a disease^19^. Additionally, there is currently a lack of an organic model explaining how genetic factors concur with COVID-19 susceptibility in patients and possibly predict the genetic severity of the disease^20^. At the time of writing this report, there is still a lack of Machine learning (ML) techniques available to experts working on Whole-exome sequencing (WES) datasets related to predicting genetic severity and providing explanations for the interaction of identified genetic variants linked to the severity of SARS-CoV-2 among patients. Explainable ML techniques that link identified genetic variant predictors can provide biological insights and interpretation that can assist experts in the field of medical sciences in understanding the complex genetic interactions that may lead to or hinder therapeutic approaches toward a trajectory to personalized medicine^21–23^. Model interpretability approaches have been adopted in several studies to discover new knowledge and testable hypotheses ^24–26^.

This study, therefore, sheds new light on genetic variants discovered from a previous study linked to the severity of COVID-19 in patients of European descent. This study provides a host genetic severity prediction and post-hoc model explanations using 16 identified candidate genetic variants and clinical covariates (age and gender) from a prior study using a 2000 cohort patients’ WES dataset^20^. The post-hoc explanation of the HGSP model predictions using the *ExplainerDashboard*. The ExplainerDashboard is an open-source python library used by social scientists to explain global or local prediction results of their model. We adopt this approach to interpret plausible complex genetic interactions that may alongside patients’ clinical covariates, interplay with the host severity of the disease outcome. The HGSP model was developed by combining several trained decision tree-based models (Random Forest and XGBoost classifiers) across a 5-fold cross-validation (CV) splitting computational strategy we employed on the original problem dataset to improve the model stability abilities ^20^. We further carried out domain interpretations of the genetic variants linked to the severity of the disease via an enrichment database (Transcription, Pathways, Ontologies, Diseases/Drugs, and Cell Types)^27^. We also linked the genetic variant via association mapping studies using the Phenome-wide Association (PheWAS) technique by leveraging an open-source bioinformatics PheWAS tool called *OpenTarget* to associate the 16 identified genetic variants with disease traits that could lead to plausible clinical trajectories of the COVID-19 disease severity in patients ^28^.

To this end, this study aimed to use the HGSP model developed from a previous 2000 cohort study of COVID-9 severity among European patients using their WES and covariates information ^20^ for out-of-sample prediction (3000 cohort WES and covariates information) and importantly, to provide post-hoc model interpretations and explanations that linked disease traits and COVID-19 severity in patients using a genetic dataset.

The rest of this paper is organized as follows: In Section 2 we explore related works, Section 3 discusses the methods, Section 4 details the methods employed, Section 5 presents the results, Section 6 discusses the results, and Section 7 concludes the study.

## 2. Related Work

Several studies have established the roles of genetic risk factors and susceptibility to COVID-19 disease, for example, the study of Choudhary & Sreenivasulu *et al*.,^8^ observed that the host genetic factors, along with other risk factors may help determine susceptibility to respiratory tract infections. It is hypothesized that the Angiotensin Converting Enzyme 2 (ACE2) gene, encoding ACE2, is a genetic risk factor for SARS-CoV-2 infection and is required by the virus to enter cells ^29^. Together with ACE2, Transmembrane Protease Serine 2 (TMPRSS2) and Dipeptidyl Peptidase-4 (DPP4) also play an important role in disease severity. According to Debnath, & Banerjee *et al*.,^4^ considering the implications of host genes in the entry and replication of SARS-CoV-2 and in mounting the host immune response, it showed that multiple genes might be crucially involved in shaping COVID-19 patients’ severity dynamics. Their study proposed three potential genetic gateways: variations within the ACE2 gene influencing the spatial transmission dynamics of the disease, the Human Leukocyte Antigen locus, a master regulator of immunity against infection which in turn seems to influence the susceptibility and severity of the disease in patients, and the genes regulating Toll-like receptor and complement pathways. This subsequently triggers the cytokine storm which induced exaggerated inflammatory pathways that seem to underlie the severity of COVID-19 in patients.

According to a study carried out by Lassau, *et al*..,^30^ observed that the SARS-CoV-2 pandemic has continued to overwhelm hospitals’ intensive care units, and the need to identify predictors of disease severity is a priority. They conducted a study on 58 clinical and biological variables, and collected a chest CT scan dataset, from 1003 coronavirus-infected patients from two French hospitals. They trained a deep learning model based on CT scans to predict SARS-Cov-2 severity in patients. Constructing a multimodal AI-severity score that includes 5 clinical and biological variables (age, sex, oxygenation, urea, platelet) coupled with the deep learning model developed an AI-severity model that improves prognosis performance. The findings of their study demonstrated that AI approaches such as neural network analysis of CT scans can bring unique prognosis information to help clinicians. The work of Muhammad *et al*.,^24^ utilizes supervised ML algorithms (Logistic Regression, Decision trees, Support Vector Machine, Naïve Bayes, and Artificial Neural Networks) using an epidemiological labeled dataset from positive and negative COVID-19 patients in Mexico. The results of their studies showed that the decision tree model outperformed all the other models in predicting the disease outcome. Yan *et al*., ^31^ utilize interpretable ML approaches to model COVID-19 patients’ mortality, and the results of their findings identified crucial predictive biomarkers of disease mortality. For this purpose, the ML tools selected three biomarkers that predict the mortality of individual patients more than 10 days in advance with more than 90% accuracy: lactic dehydrogenase (LDH), lymphocyte, and high-sensitivity C-reactive protein (hs-CRP). Relatively high levels of LDH alone seem to play a crucial role in distinguishing most cases that require immediate medical attention.

Patel, Kher, *et al*., ^12^ uses socio-demographic data, clinical data, and blood panel profile data to develop ML algorithms for predicting the need for intensive care and mechanical ventilation. The findings of their study established that the three categories of information are crucial for intensive care and mechanical ventilation allocation to patients by healthcare systems in planning for surge capacity for COVID-19. The study of Doewes, & Sharma^32^, combined an ensemble genetic algorithm and ML classifier to filter relevant features and perform classification to diagnose COVID-19 through patients’ blood samples. Their findings showed that ML techniques can augment current medical practices and tools, and improve modern methods deployed in the fight against the disease. The work of Albadr, *et al*.,^33^ employs an Optimised Genetic Algorithm-Extreme Learning Machine (OGA-ELM) with three selection criteria (i.e., random, *K*-tournament, and roulette wheel) to detect COVID-19 using X-ray images. The results from their findings showed that the OGA-ELM can be used to achieve 100.00% accuracy with fast computation time. This demonstrated that OGA-ELM is an efficient method for COVID-19 detection using chest X-ray images.

The study by Marcos, Belhassen-GarcÍa, *et al*.,^34^ demonstrated that efficient and early triage of hospitalized Covid-19 patients is crucial to detect those with a higher risk of severe Covid-19 infection. Their study trained, validated, and externally tested an ML model to early identify patients who will die or require mechanical ventilation during hospitalization from clinical and laboratory features obtained at admission. Their findings strongly supported evidence that ML models could be useful in medical emergencies such as pandemics in determining patients for hospital admission and predicting the risk of severe Covid-19 infection. Different ML techniques have been utilized by researchers in response to mitigating the SARS-Cov-2 virus threat since the onset of the pandemic some of which have been useful in the diagnosis of COVID-19 and prediction of mortality risk and severity, using readily available clinical and laboratory data^7,30^. Fallerini, *et al*.^35^ studied common and rare variants from WES data of about 4000 SARS-CoV-2-positive patients. This was utilized to define an interpretable machine-learning model for predicting COVID-19 severity. The variants were converted into separate sets of Boolean features, depending on the absence or the presence of variants in each gene. They developed an ensemble model of LASSO logistic regression which was used to identify the most informative Boolean features concerning the genetic bases of severity. Their study formed the basis of our study, however, in our study, we took a different approach to the development, interpretations, and explanations of the HGSP model. First, we used a computational strategy of a 5-fold CV to split the clinical phenotype information of the patients (i.e., each of the folds in the 5-fold CV has 80% training and 20% testing sets) and screened the 80% training sets in each fold for genetic variants (from our prior study ^19^) linked with the disease using an odd-ratio statistics approach other than the GWAS. We then remapped the identified significant variants for each of the folds for the 20% unscreened testing sets in each fold. We developed our count matrices (training and testing for each fold) following the identified variants and employed several traditional ML algorithms (Logistic Regression, Support Vector, Random Forest, and XGBoost classifiers) to identify genetic variants via feature importance from the decision tree models (i.e., Random Forest and XGBoost classifiers) that provided the highest support (i.e., non-zero feature importance weighted scores across the 5-fold). 16 genetic variants and the clinical covariates (age and gender) were identified with the highest support, and we retrain the models using these features. An ensemble voting classifier was used to combine the 5-fold decision tree models to develop the HGSP model. We validated the HGSP model on an external dataset and provided further domain interpretations and post-hoc explanations.

## 3. Materials and Methods

### Data Resources and Description

The genetic and clinical data type we used for this study was extracted from an observational study dataset coordinated by the GEN-COVID Multicenter Study group led by the University of Siena with an estimated enrolment of 2000 participants which constitutes the basis of the sample from which the genetic information was obtained (https://clinicaltrials.gov/ct2/show/NCT04549831). The initial 2000 cohort WES and covariates (age and gender) dataset were used to train and develop the HGSP model. The HGSP model was developed based on 16 identified candidate genetic variants and clinical covariates (age and gender)^20^. The out-of-sample dataset used for external prediction in this study was a follow-up dataset information provided by the same GEN-COVID group to validate the HGSP model and this formed the basis of this study. In this study, we maintained the same data preparation procedures we used for the training dataset study ^1920^. To perform the downstream analyses (external model validation and post-hoc interpretations), we considered the two classification grading schemes: adjusted-by-age and unadjusted-by-age. The adjusted-by-age grading classification scheme refers to a refinement made on the grading classification based on an ordinal logistic model which uses age as an input feature for sex-stratified patients^3,11,35^, while the unadjusted-by-age grading classification scheme was not refined. To build the feature matrices for the external model validation, we curated the allele frequencies from the WES dataset (out-of-sample) for the 16 identified genetic variants together with their clinical covariates.

We binarized the patients’ COVID-19 outcome severity grading classifications: gradings 5, 4, and 3 were grouped as “severe” and coded as 1; grading 0 was grouped as “asymptomatic” and coded as 0. We excluded patients’ information whose severity grading was classified as 1 & 2. This exclusion was done purposefully to minimize noise signals during the filtering of genetic variants that are linked with the disease severity or protection in patients. We further refined the grading classification based on an ordinal logistic model which uses age as an input feature for sex-stratified patients ^36^ and we retained only those patients whose grading classification was concordant with the one adjusted by age. The age distribution of the participants ranges from 18 years and older (Adult, Older Adult). The Gender this study considered were all sexes. Patients’ genotypic information contains a reference (Ref) or alternative (Alt) alleles in either severe or control groups which were defined by employing an additive model, whereby homozygous genotype (1/1) has twice the risk (or protection) of the heterozygous type (0/1 or 1/0).

Further details and descriptions of the WES dataset and preprocessing can be found in our prior study of the 2000 cohort dataset^20^. A total of 618 patients met the filtering and selection criteria from the out-of-sample dataset and were therefore used for the external model validation. The feature matrix for the external validation of the HGSP model contained 18 features (16 genetic variants and covariates). Each of the 16 variants that constituted the feature matrix has allelic frequency counts for each patient’s genotype information assigned (0/0 *counted as* 0, 0/1 *or* 1/0 *counted as* 1, *and* 1/1 *counted as* 2). The covariates were further merged into the feature matrix from the patients’ phenotype information. The outcome variable (grouping) is binarized as 0 and 1, where 0 represents asymptomatic and 1 depicts the severity of the disease as shown in figure 1.

**Figure 1:**
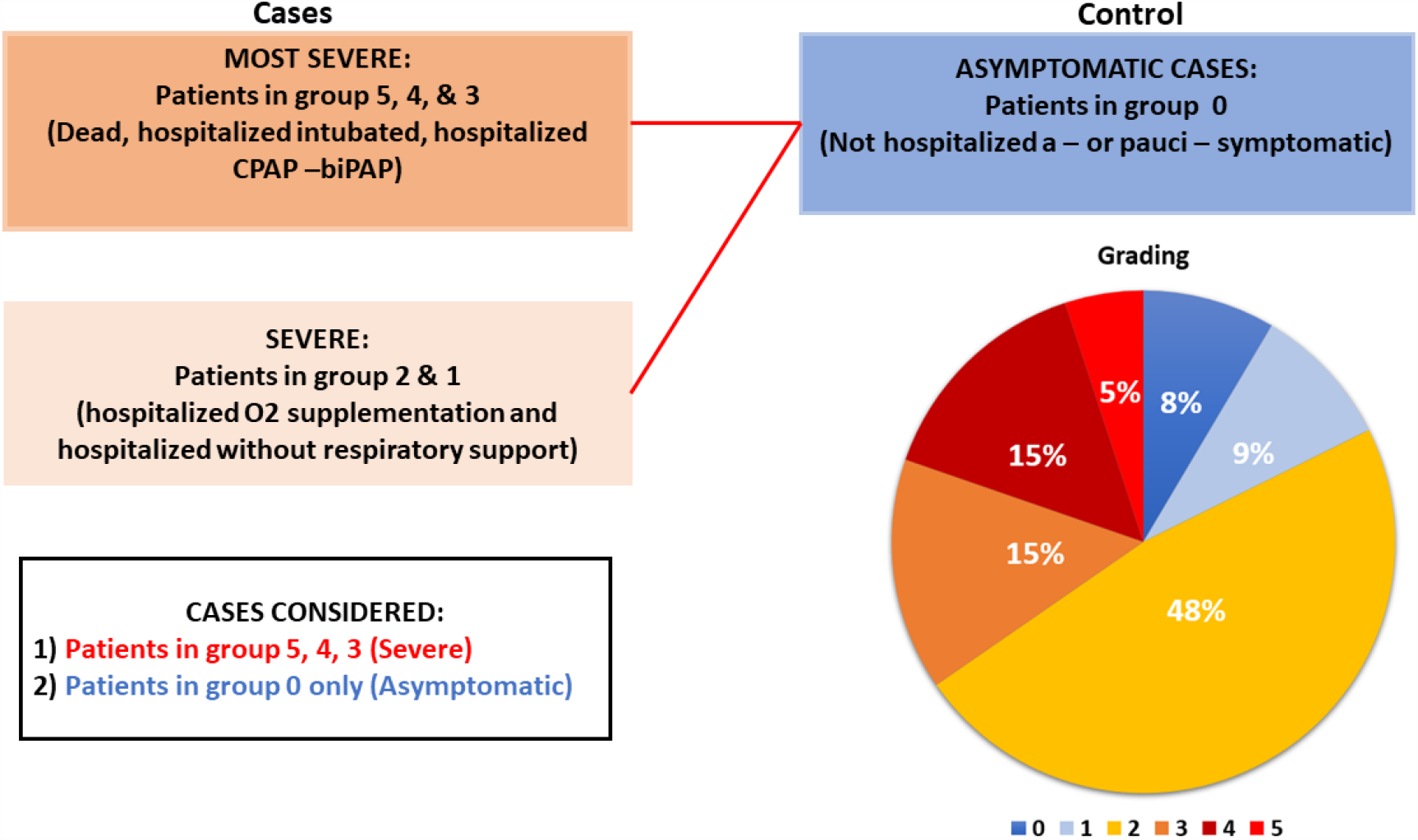
Classification of patients’ SARS-CoV-2 severity adjusted by age grading scheme. We considered the most severe cases patients in groups 3, 4, and 5 versus control cases patients in group 0.

**Figure 2:**
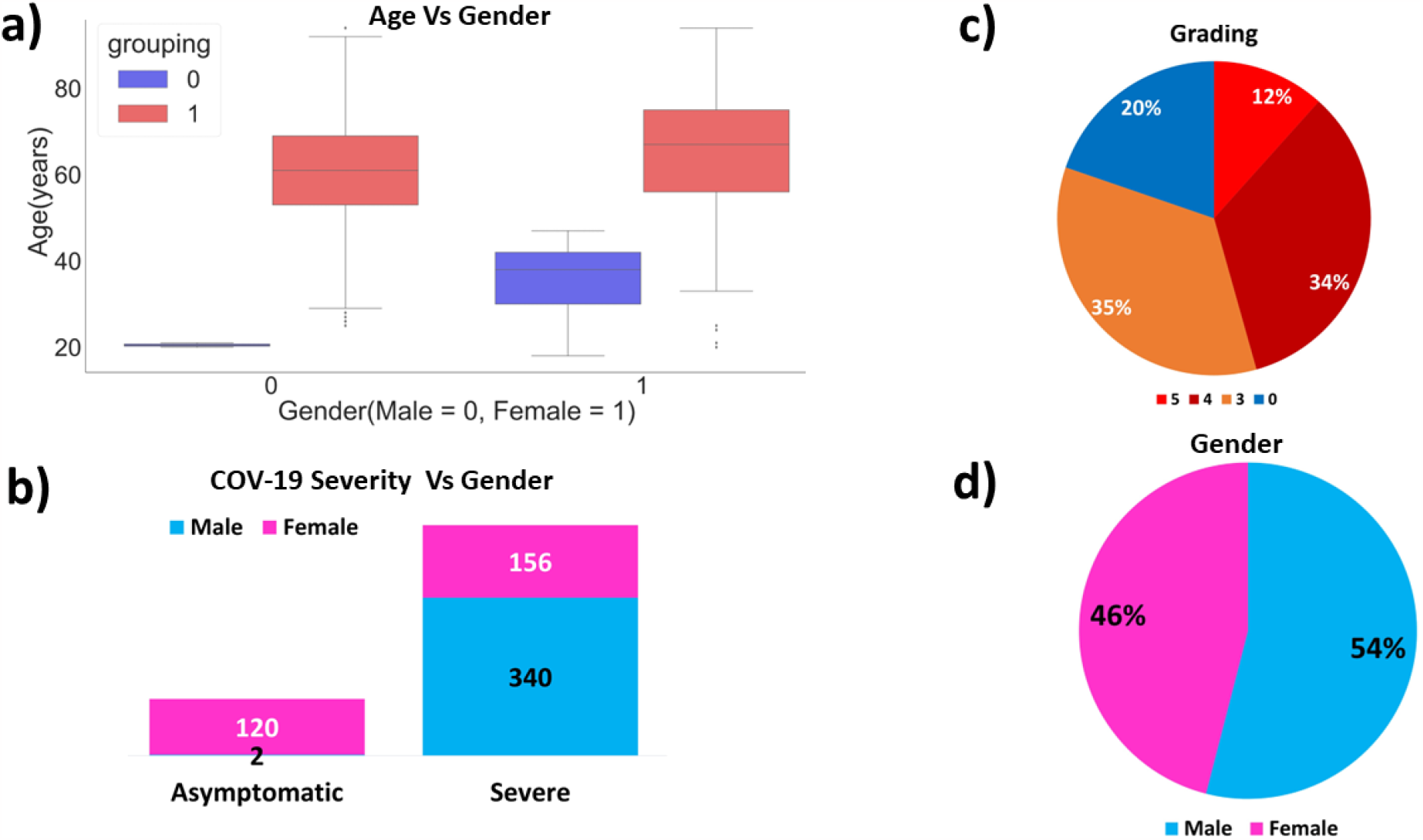
Patient phenotype information (adjusted-by-age grading scheme) in the follow-up WES dataset. We employed a four-scenario approach to validate the HGSP model using the out-of-sample dataset. The baseline scenario was mainly focused on validating the model using the adjusted-by-age grading classification scheme feature matrix. While scenarios 1 – 3 focused on validating the model using the unadjusted-by-age grading classification scheme.

### Adjusted by age grading dataset

this refers to training and testing data coming from a refinement made on the grading classification based on an ordinal logistic model which uses age as an input feature for sex-stratified patients.

### Unadjusted by age grading dataset

this refers to the training and testing data coming from an unrefined grading classification.

### Baseline training dataset

this refers to the dataset used for internal validation of the HGSP model i.e., the *20%* testing set from each of the *5-*fold CVs. There are 841 sampling units in this category and they belong to the adjusted-by-age grading scheme category.

### Testing set

this refers to the dataset used for external validation of the HGSP model, and the belongs to the adjusted-by-age grading scheme category. Note the sampling unit in this category their WES were filtered for the 16 genetic variants following the same criteria employed in the baseline training dataset. There are 618 sampling units in this category.

### Remarks

the baseline training set dataset was the initial cohort from the prior study of 2000 European descent patients WES dataset provided to us from the GEN-COVID multicenter group. They further carried out a follow-up and included additional 1000 patients making 3000 patients in all. This resulted in some overlapping and the reason for the exclusion of some samples in this study.

### Excluded samples testing set

all samples classified as severe or asymptomatic based on the unadjusted-by-age grading scheme included in the follow-up dataset. There are 235 samples in this category.

### Excluded samples training set

all samples classified as severe or asymptomatic based on the unadjusted by age grading scheme included in the baseline training dataset but excluded in the adjusted by age grading scheme baseline training dataset. There are 357 sampling units in this category.

### Aggregated excluded samples

all the samples were classified as severe or asymptomatic using the unadjusted-by-age grading scheme. They were the union of the excluded baseline training and testing samples. There are 495 sampling units in this category.

### Post-Hoc Model Explanations

For us to achieve the aim and objectives of this study, we utilized the following methodological approaches. We used the adjusted-by-age information to provide a post-hoc model explanation using the explainer dashboard framework.

### Host Genetic Severity Predictor Model Development

We developed the HGSP model by combining trained decision tree-based models (Random Forest and XGBoost classifiers) from a 5-Fold CV split of the original problem dataset. See a prior study from Onoja *et al*., ^19^ for further details. The HGSP combined these models via an ensemble “VotingClassifier” approach from the *“sklearn*.*ensemble”* python library module to aggregate the individual classifiers based on their prediction probabilities (soft margin) of the outcome.

### Prediction Metrics

The performance models were evaluated by assessing the following classification metrics; accuracy score, recall score, precision score, precision-recall curve, Matthew Correlation Coefficient (MCC), and ROC-AUC curve performance evaluation metrics to measure how well our developed covid-19 severity prediction HGSP model extrapolates the external dataset (out-of-sample dataset).

### Accuracy score

In the ML classification task, the accuracy score is a metric used to evaluate the performance of a model on a given dataset. It measures the proportion of correct predictions made by the model over all the predictions it made. In your context, the saved HGSP model was used to make predictions on an external dataset. The accuracy score was then used to evaluate how well the model performed on this dataset by calculating the proportion of correct predictions made by the model.

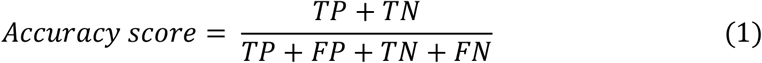

### Recall score

The recall score is a performance evaluation metric that measures the sensitivity of a model in identifying positive cases correctly. In your context, the HGSP model was validated on a new dataset using the recall score metric. This metric measures how well the model can correctly identify true positives, i.e., the number of correct positive predictions made by the model over the total number of actual positive instances in the dataset.

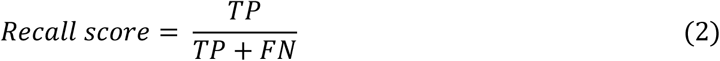

### Precision score

The precision score is a performance evaluation metric used to measure the positive predictive value of a model. It measures the proportion of true positive predictions made by the model and overall positive predictions made by the model. The precision score evaluated the performance of the HGSP model on the external validation dataset. Specifically, it was used to measure how well the model could correctly identify positive instances without producing many false positives.

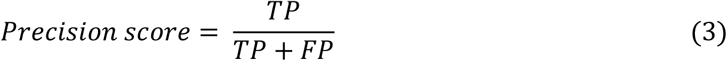

Where;

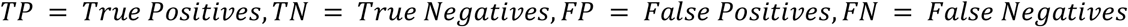

### Confusion matrix

The confusion matrix is a performance evaluation metric that shows how well a model is performing in terms of making correct and incorrect predictions on a given dataset. The confusion matrix was used to evaluate the quality of the output of the HGSP voting classifier on the dataset. The confusion matrix is a square matrix that displays the number of correct and incorrect predictions made by the model across distinct categories or labels. The diagonal elements of the matrix represent the number of points for which the predicted label is equal to the true label, while the off-diagonal elements represent those that were mislabeled by the classifier.

### f1-score

The f1-score is a performance evaluation metric that measures the model’s accuracy in binary classification tasks. The f1-score is the harmonic mean of precision and recall. The f1-score ranges from 0 to 1, where 1 represents perfect precision and recall, while 0 represents the worst possible performance. The f1-score is often used as a benchmark to compare the performance of different binary classification models.

### Precision-Recall Curve

The Precision-Recall (PR) curve is a performance evaluation metric that is commonly used to assess the quality of a binary classification model. The precision-recall curve is generated by plotting the precision and recall values of a binary classifier for different probability thresholds.

### Matthew Correlation Coefficient

The MCC is a performance evaluation metric that measures the quality of binary classification models. MCC is a value between -1 and +1, where +1 represents perfect classification, 0 represents random classification, and -1 represents the worst classification. MCC considers all four elements of the confusion matrix (True Positive, True Negative, False Positive, and False Negative) and is therefore considered to be a more robust measure than other binary classification metrics, such as accuracy or F1 score.

The MCC is calculated using the formula:

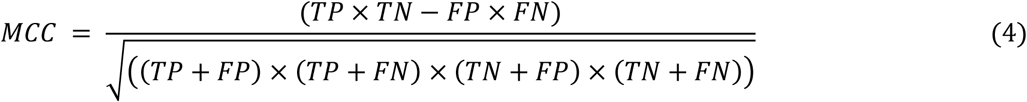

where TP, TN, FP, and FN represent the number of true positives, true negatives, false positives, and false negatives, respectively.

### Log-loss

This is a crucial classification metric we used to assess the performance of the ensemble voting classifier. Log-loss is indicative of how close the prediction probability is to the corresponding actual/true value (0 or 1 i.e., binary classifier). The more the predicted probability diverges from the actual value, the higher the log-loss value.

### ROC Curve

The Receiver Operator Characteristic *(ROC)* curve is a graphical representation plot that is used to show the diagnostic ability of binary classification tasks. The ROC curve was used to visualize how well the saved HGSP model used for the study extrapolates the external dataset ^37^.

### Interpreting the ROC curve

The ROC curve was used to show the trade-off between sensitivity (True Positive Rate *(TPR)*) and specificity *(1 – FPR)* of the HGSP model. If the voting classifier produces curves closer to the top-left corner indicates a better performance. For baseline purposes, a random classifier is expected to give points lying along the diagonal *(FPR = TPR)*. The closer the curve comes to the 45-degree diagonal of the ROC space, the less accurate the test could be.

### ExplainerDashBoard Visualization

This is an open-source python package ^38^ that makes it convenient to quickly deploy a dashboard web app that explains the inner workings of a (scikit-learn compatible) machine learning model. The dashboard provides interactive plots on model performance, SHAP feature importance, feature contributions to individual predictions, “what if” analysis, partial dependence plots, feature importance, SHAP (interaction) values, visualization of individual decision trees, etc.

### SHAP Feature Importance

The ExplainerDashboard python library has an inherent feature importance criterion built into the explainer model ^39,40^. This helps to calculate a score for all the 16 fully supported variants and covariates (age and gender) input features and is displayed as bar plots. In this context, the scores represent the “importance” of each feature. A higher score means that the specific feature has a larger effect on the model that is being used to predict a COVID-19 genetic severity. In this study, we further investigated the SHAP feature importance output from the explainer dashboard by visualizing complex host genetic interactions with covariates via a clustering heatmap.

## Domain Interpretation

### Phenowide Disease-Variants Association studies

The phenome-wide Association studies (PheWAS) is an approach we used to analyze many phenotypes to compare with the genetic variants we used for the supervised ML modeling. By utilizing comprehensive Genome information (DNA dataset) one can link the disease status or other traits of an individual such as disease complications or adverse drug events. In this study, we used the **OpenTarget web tool** for a disease-variant association, and the **PheWAS** p-value cut-off was set to (α < 0.001)^28^. We considered only positive **PheWAS** association (odds-ratio > 1) where the trait was associated with severe cases and not asymptomatic (case > 1). We focused on the 16 genetic variants used to develop the HGSP model for external prediction. The PheWAS profile information for each of the 16 variants was integrated into the “explainerdashboard” SHAP feature importance bar plots and SHAP feature importance heatmap visualizations of the prediction outcome.

The Enrichr is a web-based interactive software tool that integrates various gene-set libraries and provides alternative approaches to rank enriched terms. It also includes interactive visualization methods to display enrichment results using the D3 library. Enrichr can be embedded into gene list analysis tools to provide domain knowledge interpretations such as Transcription, Pathways, Ontologies, Diseases/Drugs, and Cell Types. In this case, Enrichr was used to further link the 16 genetic variants used in the HGSP model for biological implications.

## 4. Results

We present results from a validation of the trained HGSP model we developed using the 16 identified genetic variants and clinical covariates (age and gender) considering the adjusted-by-age grading scheme (see Fig. 1). We further provided post-hoc model interpretations for the HGSP model prediction outcome considering the unadjusted-by-age grading scheme.

### External Model Validation

We consider the following case studies for model validation using the 16 identified candidate genetic variants and two clinical covariates (age and gender). 1) Baseline training dataset, 2) Testing set, 3) Excluded samples Testing set, 4) Excluded samples Training set, 5) Aggregated excluded samples,

In this study, we utilized the HGSP model we developed from a prior study of training decision tree-based models (Random Forest and XGBoost classifiers) combined from across a 5-fold CV (see further details in Onoja et al., ^19^) The HGSP model voting classifier was developed from high-performance machine learning algorithms that have some interpretability abilities due to their recursive tree-based decision system. We used this approach rather than adopting a complex model such as a deep neural network model to minimize the risk of overfitting and avoid the black box approximation of the problem considering their internal model mechanisms are difficult to interpret. The model explanation was further done using the explainer dashboard approach to assess and visualize vital metrics at a local level such as the SHAP permutation feature importance and dependence plots. The SHAP feature importance weight in our ensemble model is determined by its accumulated use in each decision step and permutation of each feature, shuffled and sorted based on their absolute values. Visualizing the 18 features via the bar plot helps us to further understand the relative importance of each of the 18 features in estimating the most discriminative COVID-19 outcome severity prediction in patients. Table 1 summarizes the performance of the ensemble model on the external validation set.

**Table 1:** Summary of Performance Metrics external model validation of all case studies

| Case study | Accuracy | f1-score | Precision | Recall | MCC |
| --- | --- | --- | --- | --- | --- |
| Testing set | 82.85 | 88.09 | 99.49 | 79.03 | 64.08 |
| Excluded samples Testing set | 83.83 | 88.82 | 95.57 | 82.97 | 62.12 |
| Excluded samples Training set | 82.35 | 85.52 | 88.15 | 83.04 | 63.17 |
| Aggregated excluded samples | 84.44 | 88.14 | 90.51 | 85.89 | 65.79 |

The results show that our developed HGSP model voting classifier accurately recapitulates vital information that predicts the severity outcome in COVID-19 patients, regardless of their cohort. It is also worth noting that the performance of the external dataset is similar to that of the training and validation sets obtained in the training cohort dataset. This implies that the HGSP model voting classifier can capture the key relevant information that predicts patients’ severity outcome of COVID-19 disease.

### Results of Analysis Post-hoc HGSP Model Interpretations and Explanation

Next, we performed the post-hoc model agnostic interpretations and explanations at a local level for the HGSP predictions. We employed the ExplainerDashboard interpretation and explanation approach focusing on the SHAP dependence plots, and feature importance plots, results to further shed new light on understanding complex interactions of genetics with clinical covariates. Here we seek to unravel hidden insights such as patients whose COVID-19 severity predictions are not driven by covariates (age and gender) but as a result of some complex genetic interactions from the 16 identified consistent features. The HGSP does not perform basic EDA approaches such as descriptive statistics summary and bar or histogram plots. The ExplainerDashboard provides options to view different post-hoc interpretation results. The ExplainerDashboard results can be saved in PDF, HTML, JPEG, or PNG file formats. Displayed in Figure 5(a) – 5(e) are the results of the HGSP model from the ExplainerDashboard interactive interface.

**Figure 3:**
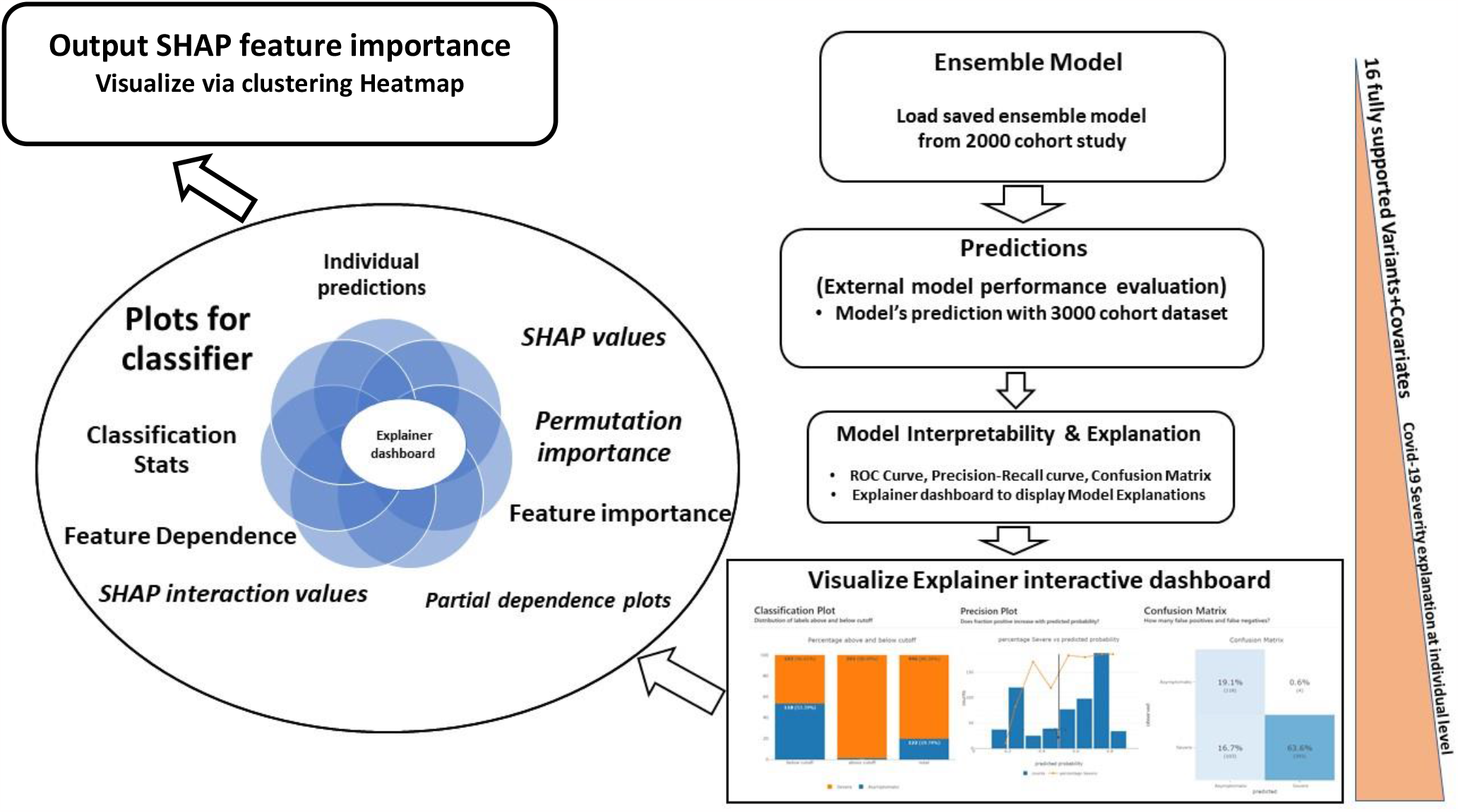
Proposed methodological design of COVID-19 HGSP model post-hoc explanations. We first load the saved HGSP model developed by training fully supported variants and covariates (age and gender) identified from a simple stratified 5-fold CV splitting strategy adopted from the 2000 cohort dataset study. We proceeded to validate the model on the out-of-sample dataset. Our method used the **HGSP** for prediction purposes and employed the **ExplainerDashboard** python library to provide a post-hoc model explanation of the prediction outcome at an individualistic level using the adjusted by-age information (baseline scenario). Note full details on how the HGSP model was developed and trained are provided in a study by Onoja *et al*. ^19^.

**Figure 4:**
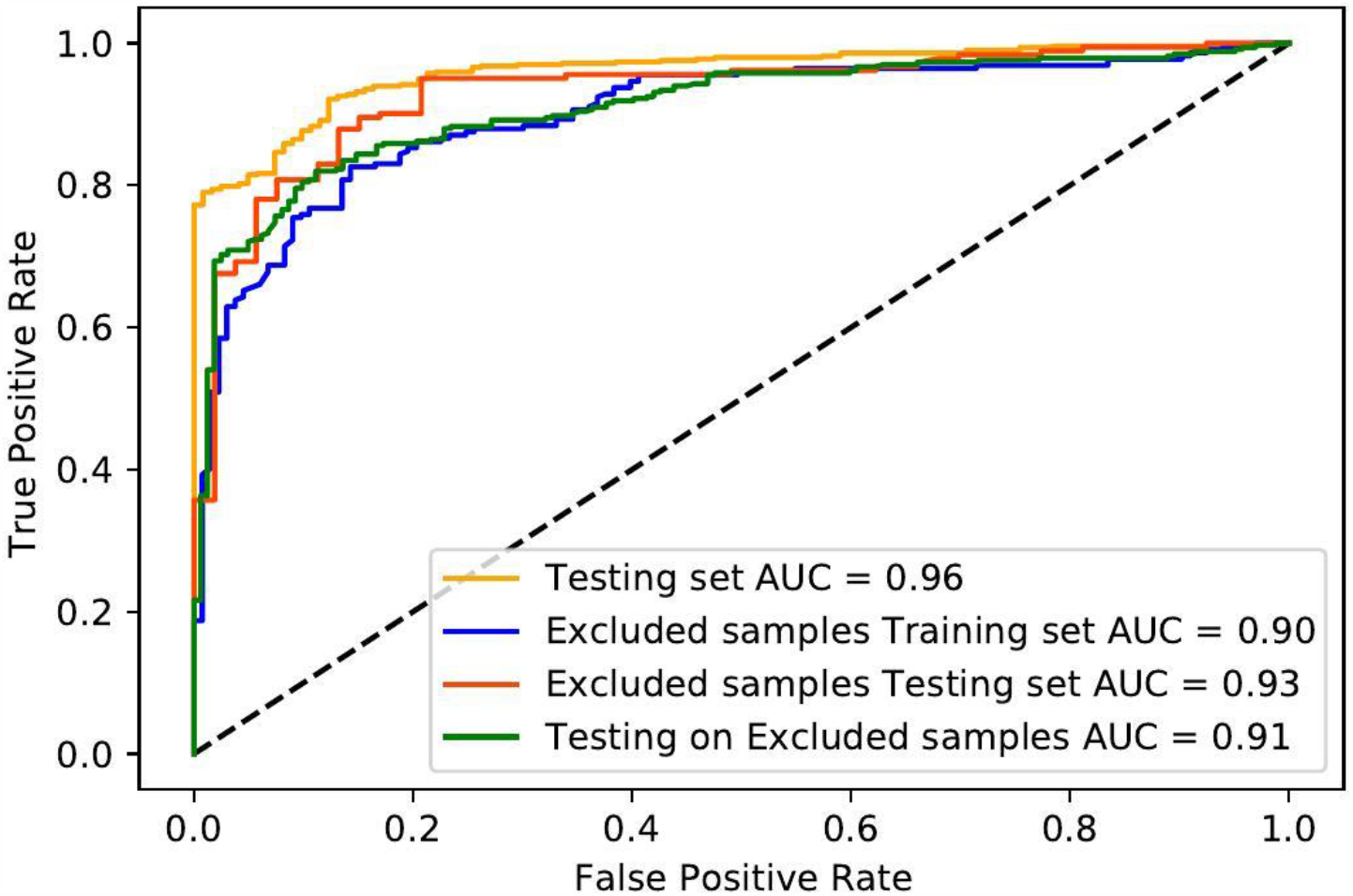
HGSP model performance considering out-of-sample model validation in all case studies.

**Figure 5(a):**
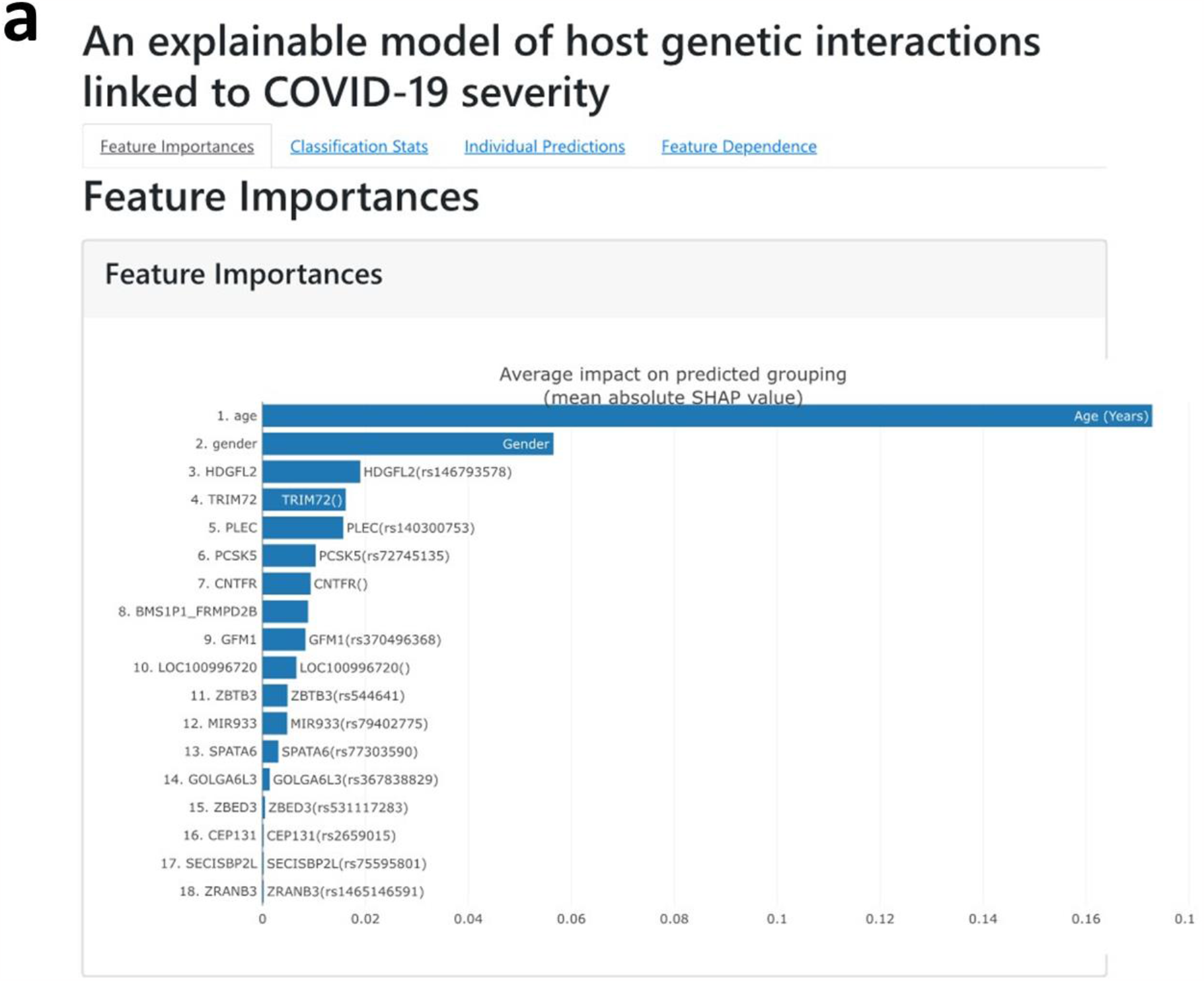
ExplainerDashboard displayed SHAP feature importance plot. The plot shows the features sorted from most important to least important. The features were sorted based on the absolute SHAP values (average absolute impact of the 18 features on the final prediction outcome). The features can also be shuffled and sorted based on their permutation importance.

**Figure 5(b):**
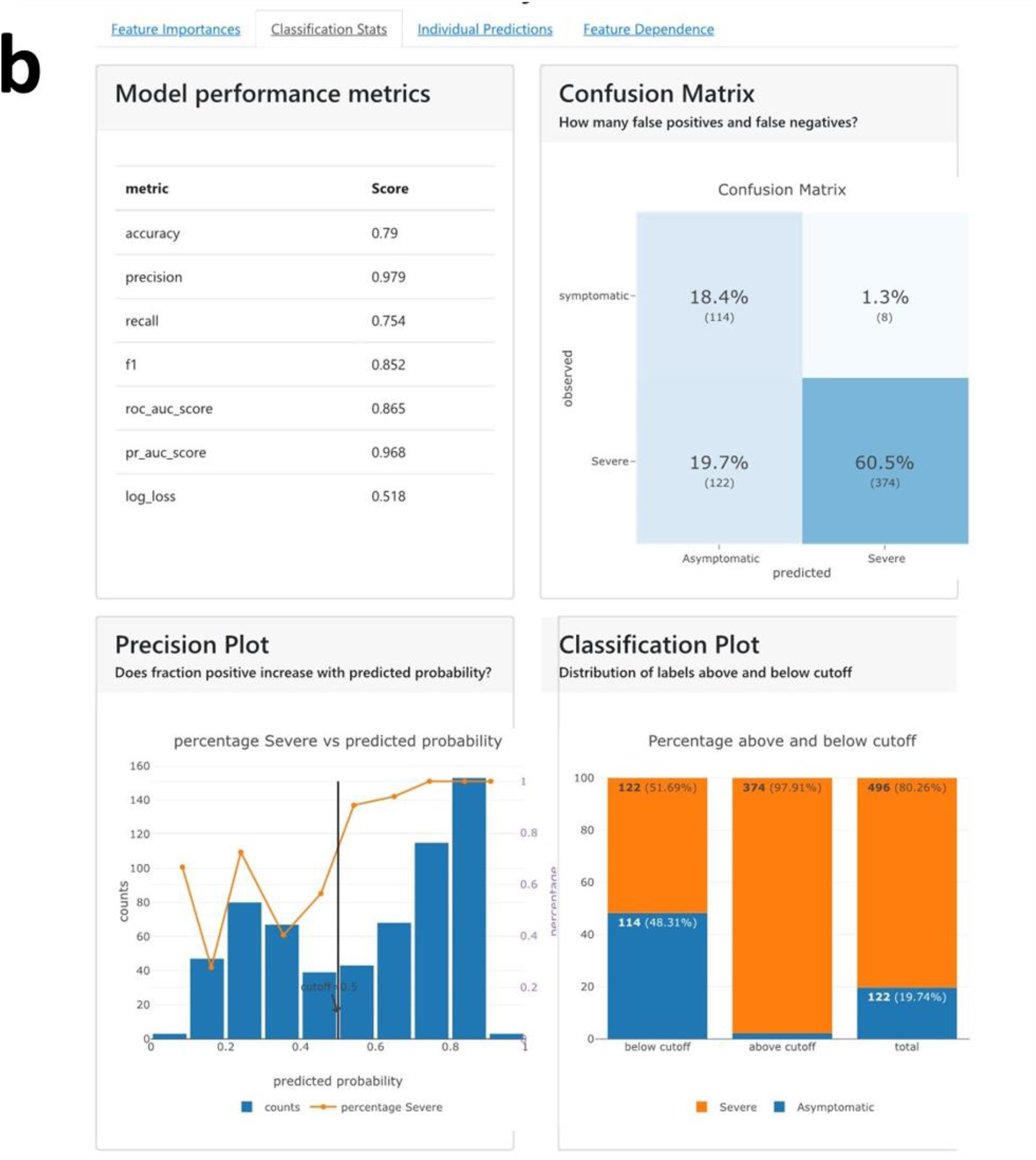
ExplainerDashboard displayed Classification Stats. In figure 5(b) we show the ExplainerDashboard displayed the classification statistics results of the HGSP prediction: confusion matrix plot; precision plot and classification plot. The observations were binned together in a group of roughly equal predicted probabilities and the percentage of positives is calculated for each bin. A perfectly calibrated model would show a straight line from the bottom left corner to the right corner. A strong model would classify most observations correctly and close to *0%* or *100%* probability the classification plot displayed the fraction of each class above and below the probability cut-off of 0.50: the ROC curve performance of the ensemble model on an external prediction dataset.

**Figure 5(c):**
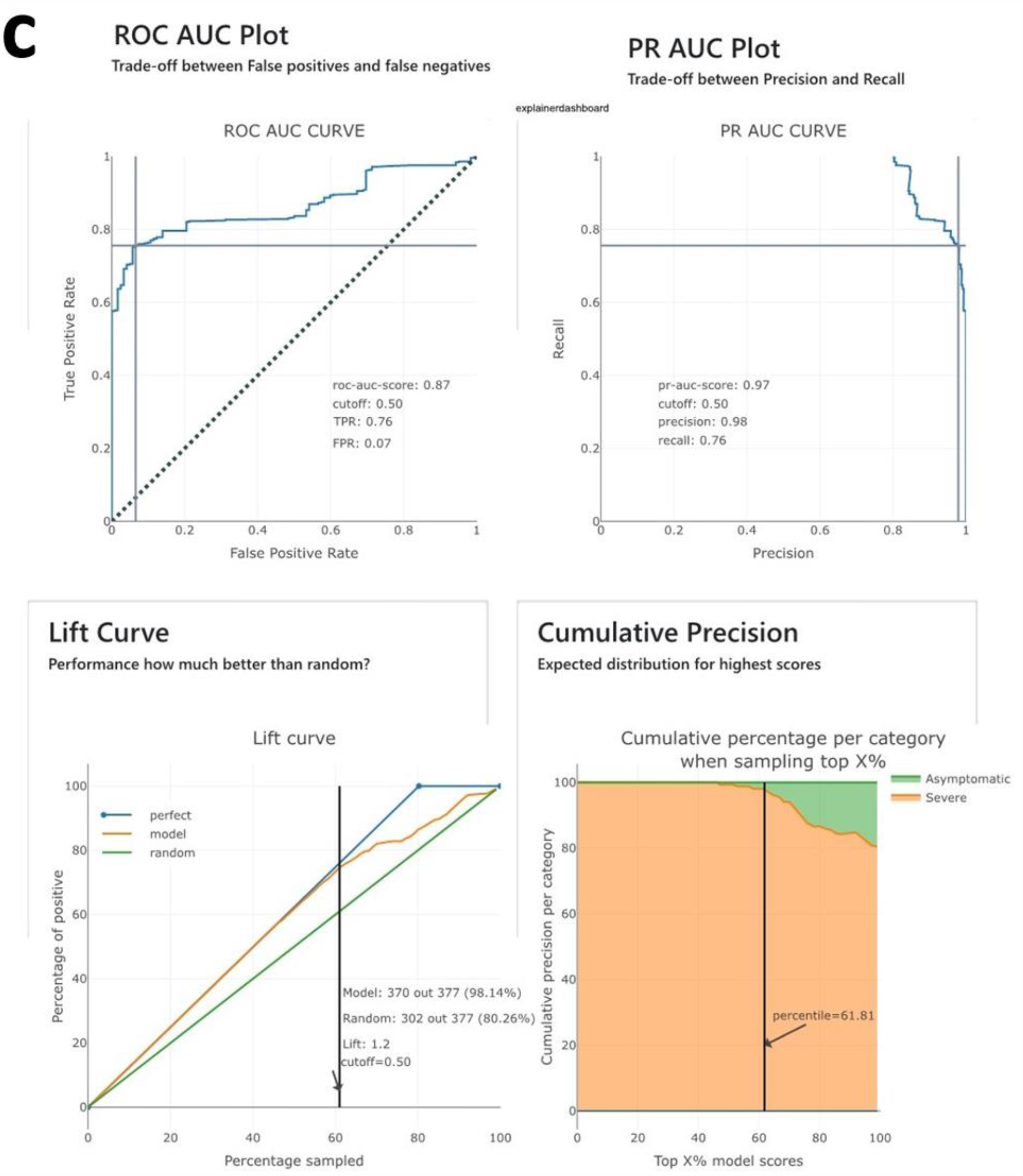
ExplainerDashboard displayed Classification Stats. The plots displayed the Precision-Recall Area Under Curve (AUC) performance on an external follow-up cohort dataset; the lift curve plot is used to depict the percentage of positive classes when one selects only observations with a score above the cut-off Vs selecting the observations randomly. It aimed to help us evaluate how much better our developed ensemble voting classifier is than a random (the lift).

**Figure 5(d):**
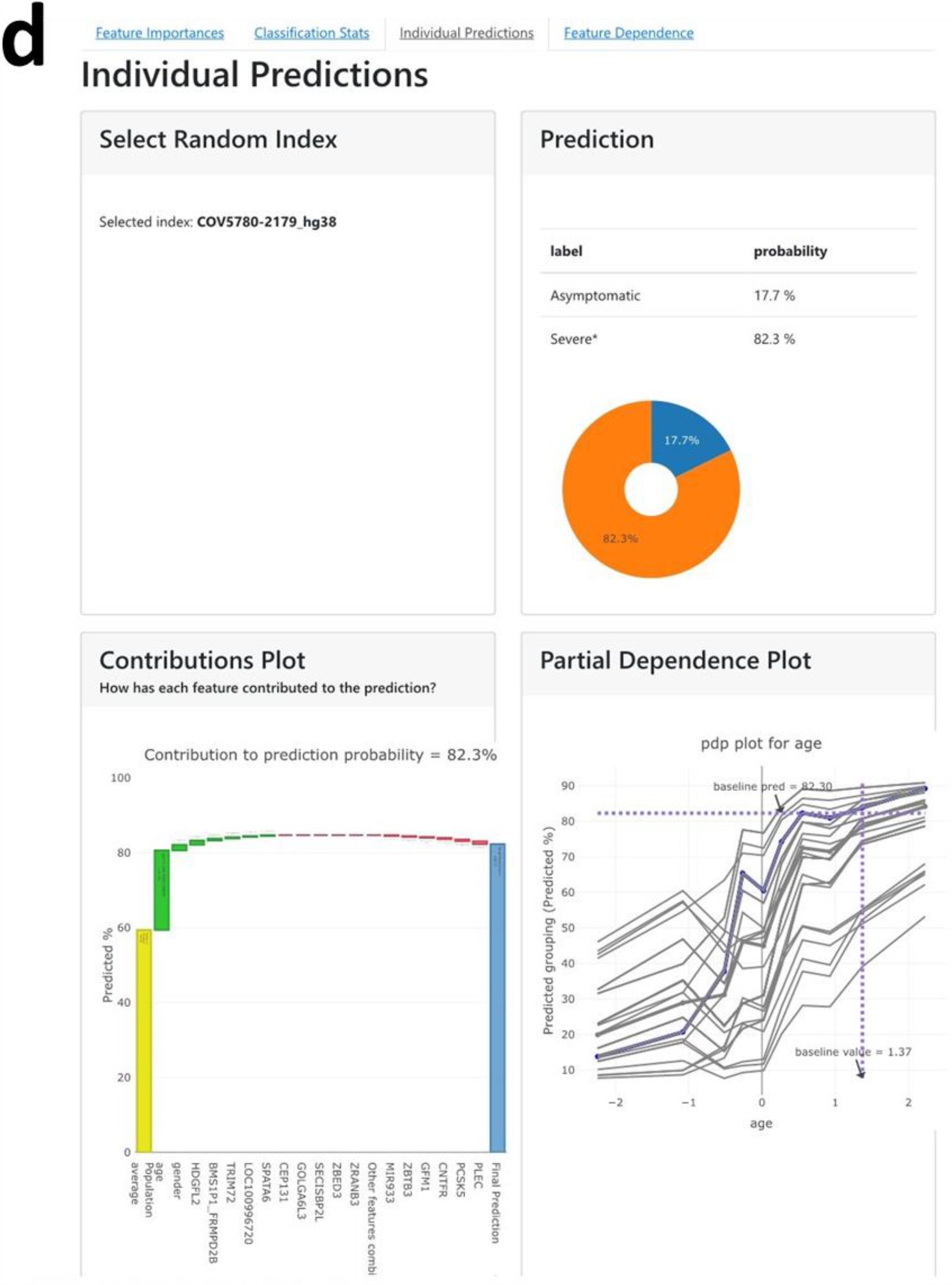
ExplainerDashboard Individual predictions plot. The dialogue box is pulled down to select a sample_ID directly by choosing it from the dropdown list or hitting the random sample_ID button to randomly select a sample_ID that fits the constraints. This aimed to help us assess in general the false positives and false-negative rates of our prediction. The doughnut prediction plot shows the predicted probability for each grouping label for the selected sample_ID of interest.

**Figure 5(e):**
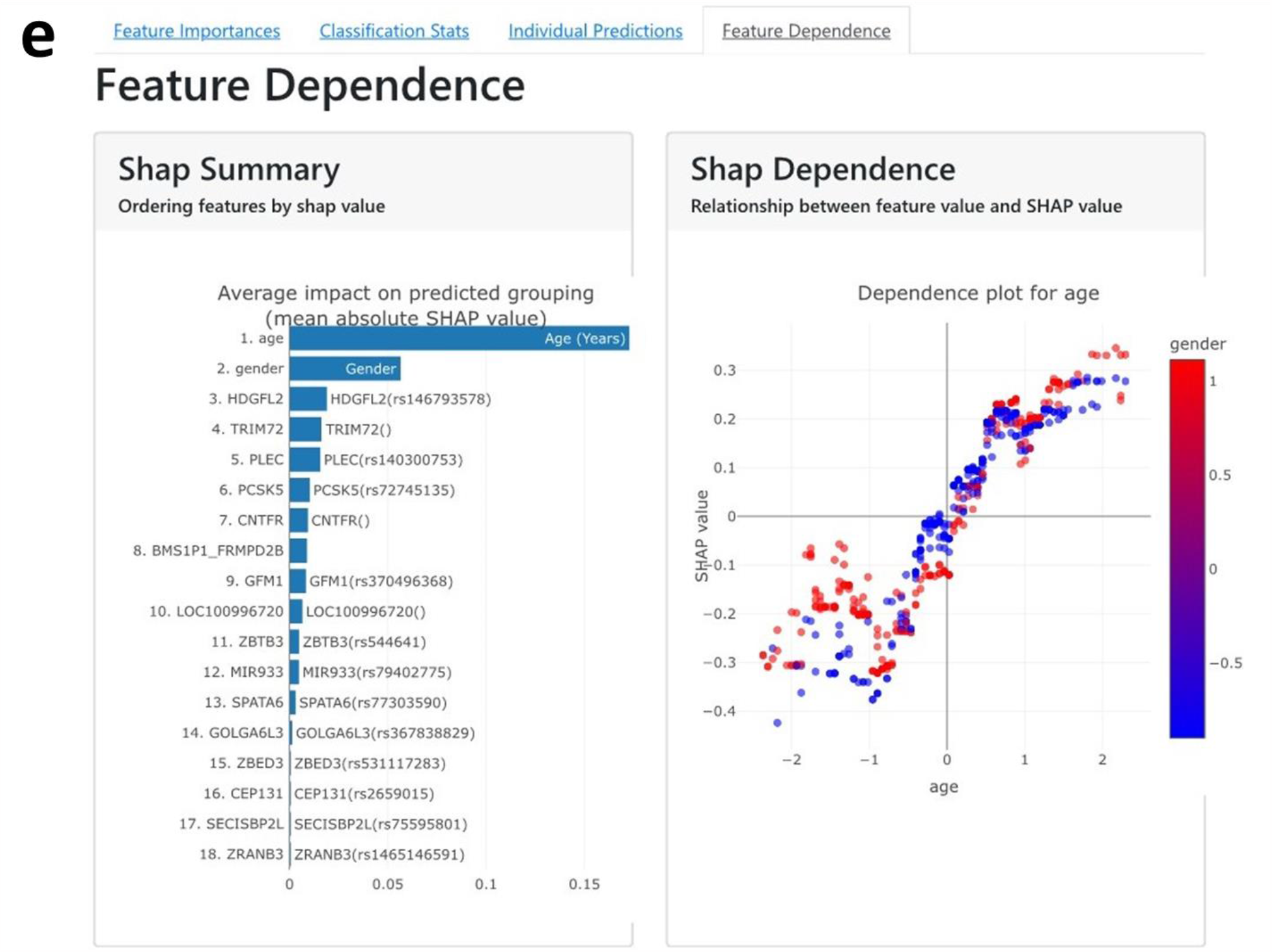
ExplainerDashboard Feature dependence plot. The SHAP dependence plot displays the relationship between feature values and SHAP values. This aimed to allow us to investigate the general relationship between feature value and impact on the prediction. One can ascertain whether the model uses features as expected or uses the plots to learn more about the relationships that the model has learned between the input feature and the predicted outcome.

### Domain Interpretations and Explanations

We linked the genetic variants used to validate the HGSP model and ExplainerDashboard with the OpenTarget genetics and Enrichr bioinformatic web-based tools. The PheWAS results from OpenTarget genetics are presented in table 2 while Figure 6 presented a snapshot interface of the Enrichr results for the top 15 genetic variants.

**Table 2:** Feature importance description associated with PheWAS analysis

| *Feature* | *Description* |
| --- | --- |
| *gender* | Gender |
| *age* | Age (Years) |
| *HDGFL2* | HDGFL2(rs146793578) [**Hint** - cardiovascular disease: Hypertension, Phenotype: Fasciitis, Cell proliferation disorder: Prostate cancer\| illnesses of siblings] |
| *TRIM72* | TRIM72 ( ) |
| *PLEC* | PLEC (rs140300753) [**Hint** - Phenotype: Abnormalities of breathing, Cardiovascular disease: Heart attacks] |
| *PCSK5* | PCSK5 (rs72745135) [**Hint** - Phenotype: Abnormalities of breathing, Mouth breathing, Cardiovascular disease: Epistaxis or throat haemorrhage, Infectious disease: Other acute lower respiratory infections] |
| *CNTFR* | CNTFR ( ) |
| *BMS1P1_FRMPD2B* | BMS1P1_FRMPD2B ( ) |
| *GFM1* | GFM1(rs370496368) ( ) |
| *LOC100996720* | LOC100996720 ( ) |
| *ZBETB3* | ZBETB3 (rs544641) [**Hint** - Infectious disease: Viral hepatitis] |
| *MIR933* | MIR933 (rs79402775) |
| *SPATA6* | SPATA6 (rs77303590) [Hint - Immune system disease: Autoimmune disease, Infectious disease: Infectious mononucleosis / glandular fever / epstein barr virus (ebv), Viral hepatitis, Cardiovascular disease: Hypertension] |
| *GOLGA6L3* | GOLGA6L3(rs367838829) ( ) |
| *ZBED3* | ZBED3 (rs531117283) [**Hint** - Biological process: Frequent intake of alcohol, Infectious disease: Meningitis \| non-cancer illness code] |
| *CEP131* | CEP131 (rs2659015) [**Hint** - Biological process: Current smoking status, Cardiovascular disease: Esophageal bleeding (varices/haemorrhage)] |
| *SECISBP2L* | SECISBP2L(rs75595801 ) [**Hint** - Genetic, familial or congenital disease: Disorder of lipoprotein metabolism, Phenotype: Haemorrhage from gastrointestinal ulcer] |
| *ZRANB3* | ZRANB3(rs1465146591) |

**Figure 6:**
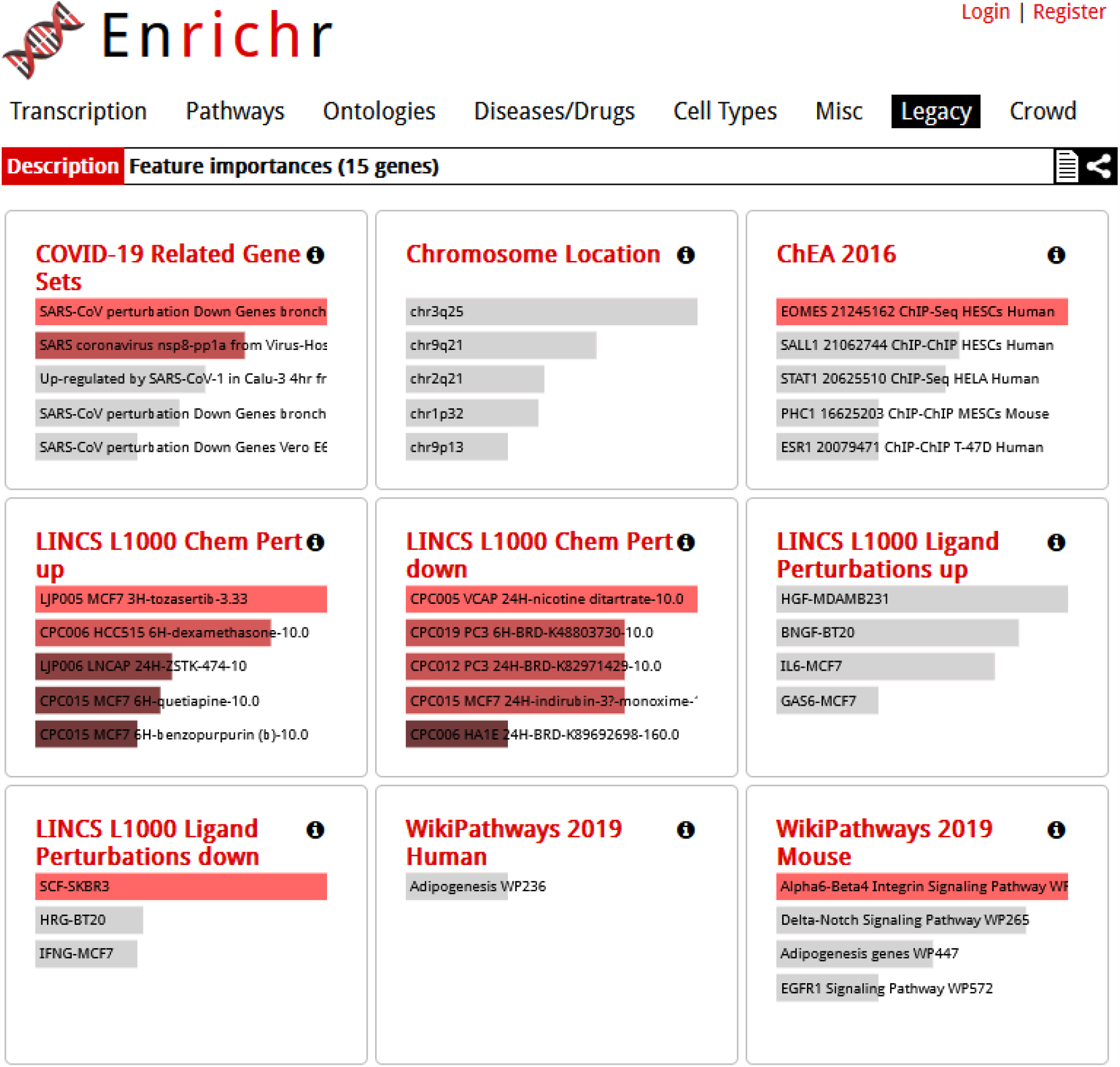
Snapshot of Enrichr web-based results linking the genetic variants used for the HGSP Model. The detailed results of the Enrichr domain interpretations can be found using the link below: https://maayanlab.cloud/Enrichr/enrich?dataset=26f3365c99e0255115dd818c11aba294#

Next, we presented the results of the SHAP value feature importance from the ExplainerDashboard visualizing it via the hierarchical clustering heatmap (See Fig. 7).

**Figure 7:**
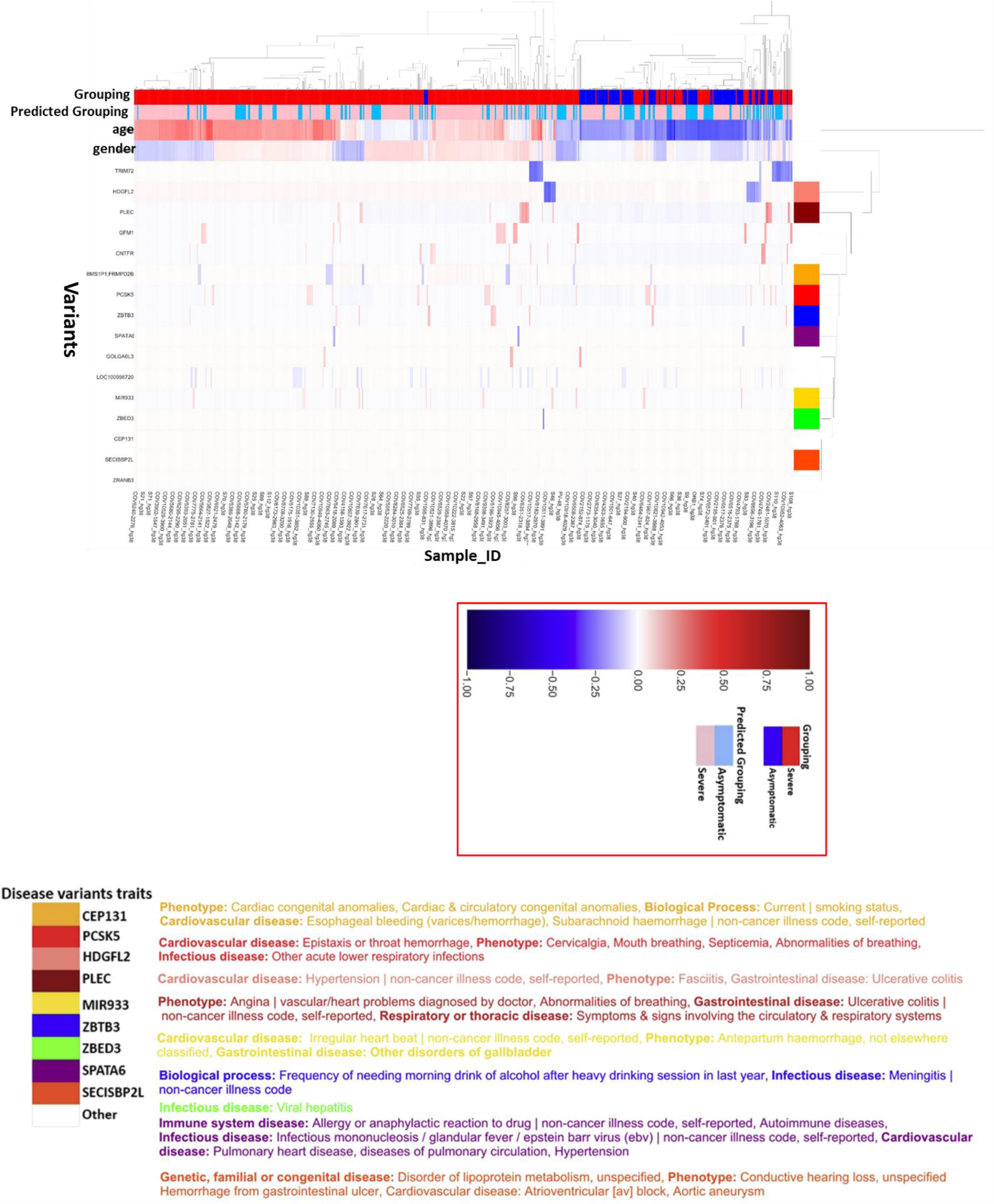
ExplainerDashboard clustering heatmap visualizations of SHAP features importance output. The covariates and variants (Fully supported variants) at the local explanations level we visualized the SHAP feature importance output for plausible interactions interplaying with COVID-19 severity predictions. The Shapley values for each feature’s important interpretations and explanations range from negative to positive. Negative values were colored blue while positive were colored red. A positive value means the feature is pushing the prediction output in a forward or positive direction while a negative value means the feature is pushing the output backward. Meaning features with positive pull force will favor grouping 1 (severe) while features with backward pull force (negative) will favor grouping 0 (asymptomatic).

## 5. Discussion

There is still a lot we are yet to unravel when it comes to the severity manifested by different SARS-CoV-2 patients. For example, why are certain patients even though not advanced in age and with no comorbidity susceptible severity to the disease while others are not? The significance of our study is three folds, first, we identified 16 candidate genetic variants that are likely determinants of COVID-19 severity outcomes in patients from a prior study we carried out using the 2000 cohort WES and clinical datasets. We further developed by combining several traditional interpretable ML algorithm models (decision tree-based models – Random Forest and XGBoost classifiers across a simple stratified trained 5-fold split CVs) to form the HGSP model and carried out an external validation using a follow-up dataset. Secondly, we utilized the explainer dashboard open-source python library to carry out a post-hoc explanation of our model predictions particularly considering the out-of-sample dataset. We employed the domain knowledge of the Phenome-wide Association technique by leveraging the OpenTarget web tool to associate the 16 identified genetic variants with disease traits that could lead to plausible clinical trajectories of the COVID-19 disease in patients. Lastly, we used the hierarchical clustering visualization heatmap to unravel hidden insights from the SHAP feature importance values from the explainerdashboard.

From the heatmap (see Fig. 7) the covariates of age and gender were separated along two lines of magnitude impacts (positive and negative directions). This means they can be a push forward (severe) or pull back (asymptomatic) to the model severity prediction. This is in line with existing findings from the literature that the male gender is more at risk of severe COVID-19 disease than the female gender. More so, research has shown that as one advance in age, they are more likely at risk of severe COVID-19 than younger patients.

Visualizing the SHAP value feature importance via a hierarchically clustered heatmap helps us to further understand the directionality contributions of the features at an individualistic level for each patient’s severity predictions. For example, the variants PLEC & PCSK5 (see Fig. 7) are strongly associated with abnormalities of breathing, symptoms, and signs involving the circulatory and respiratory systems. Most of the patients’ genetic severity prediction is strongly linked to age and gender, however, there is a small portion of patients whose severity predictions are topmost contributed by genetic variants; for example, patients with sample IDs S46_hg38, COV3908-1549_hg38, COV5958-2221_hg38, COV6351-2318_hg38, COV6807-2447_hg38, COV7658-2744_hg38, COV7878-2817_hg38 and COV8603-3159_hg38, whose severity contribution is coming from the variant PLEC, and not gender or age covariates. The variant PLEC has been linked with abnormalities in breathing phenotype traits and may be a suspected candidate disease variant whose interactions may interplay with the disease outcome in the identified patients. This implies that not all the patients’ severity is purely driven by the covariates’ age and gender, some of which come from the genetic interactions of the host with the disease. This is in line with our previous study which utilizes the Principal Component Analysis (PCA) on the training dataset and identified a small cluster group of patients (about 29 in number with 98% homogeneous COVID-19 severity). When these patients’ gene lists were further investigated for biological implications via the pathway enrichment, biological processes such as the JAK-STAT signaling pathway, Cytokine-cytokine receptor interaction, and Interleukin-6 family signaling were detected. This revealed that genetics may have inter-play with the severity of the SARS-CoV-2 virus of patients in this cluster grouping.

Also, the HGSP model is further deployed as a web application to assist experts working with WES datasets of COVID-19 patients and seek to evaluate their model based on the 16 identified genetic variants and clinical covariates. For example, an individual patient (see Fig. 5) can zoom in to understand how genetics is interacting with clinical covariates in predicting the severity outcome of the disease. The patient even though young and possibly healthy could be at susceptible risk to the disease if exposed to the virus. However, it is worth noting that the HGSP model tends to favor severity outcome predictions compared to the asymptomatic cases, partially due to inherent imbalanced class distributions in the problem dataset. This reflects the reality that during the pandemic’s first and second waves, there was a high surge of patients infected by the disease which led to a high influx of admitted severe patients and the collapse of healthcare facilities in many countries. The HGSP model, therefore, could serve as a good decision tool to lessen healthcare cost burdens by decision-makers to identify patients with true COVID-19 severity. Also, the HGSP model can be used to explain the rationale behind the patients’ severity prediction outcomes as to why they were selected or not. Developing a user-friendly explanation ML system can augment the efforts of healthcare decision-makers and clinicians to further build trustworthy and explainable models that will promote efforts toward personalized medicine.

## 6. Conclusion

Providing explanations and insights to the 16 identified genetic candidate variants we used to develop the HGSP model and validated it on an external follow-up dataset is crucial and formed the major contribution of this study. The findings from this study shed more insights into our understanding of the complex genetic interactions that the 16 identified genetic variants may be interplaying in the host genetic severity of COVID-19 disease. Most of the patients’ severity has been contributed by age, gender covariates, and other modulated factors such as comorbidities, however, there is a small portion of the patients whose severity is purely driven by genetics. This group of individual patients spotted in the population might be at unusually high risk and need to be protected against the SARS-CoV-2 infection. In the future, we seek to investigate the WES patients’ dataset using statistical and bioinformatic tools such as linear regression SKAT-O analysis, polygenic variant scoring, and deep neural knowledge prior networks to provide domain experts with biologically inspired interpretability abilities for robust data-driven solutions.

## Data Availability

The cleansed dataset and codes are available on our Githhub group page at: https://github.com/raimondilab/COVID-19-severity-host-genetic-predictor-model-explanation

## Data and code availability

We confirmed that all materials used for this study are readily available from the authors upon request. Further information, sample feature count matrix, codes, and figures are available in the Supplementary Information. The code implementation is available at https://github.com/raimondilab/COVID-19-severity-host-genetic-predictor-model-explanation Enrichr results: https://maayanlab.cloud/Enrichr/enrich?dataset=26f3365c99e0255115dd818c11aba294#

## Acknowledgments

We dedicate this paper to all those who have devoted their time, resources, and lives to waging embittered battle against the raging coronavirus pandemic.

## References

1. Zhu, N. et al. A Novel Coronavirus from Patients with Pneumonia in China, 2019. New England Journal of Medicine 382, 727–733 (2020).

2. Gursel, M. & Gursel, I. Is global BCG vaccination-induced trained immunity relevant to the progression of SARS-CoV-2 pandemic? Allergy: European Journal of Allergy and Clinical Immunology 75, 1815–1819 (2020).

3. Fallerini, C. et al. Association of toll-like receptor 7 variants with life-threatening COVID-19 disease in males: Findings from a nested case-control study. Elife 10, (2021).

4. Debnath, M., Banerjee, M. & Berk, M. Genetic gateways to COVID-19 infection: Implications for risk, severity, and outcomes. The FASEB Journal 34, 8787–8795 (2020).

5. Alballa, N. & Al-Turaiki, I. Machine learning approaches in COVID-19 diagnosis, mortality, and severity risk prediction: A review. Inform Med Unlocked 24, 100564 (2021).

6. Duckworth, C. et al. Using explainable machine learning to characterise data drift and detect emergent health risks for emergency department admissions during COVID-19. Scientific Reports 2021 11:1 11, 1–10 (2021).

7. Liang, W. et al. Development and validation of a clinical risk score to predict the occurrence of critical illness in hospitalized patients with COVID-19. JAMA Intern Med 180, 1081–1089 (2020).

8. Choudhary, S., Sreenivasulu, K., Mitra, P., Misra, S. & Sharma, P. Role of Genetic Variants and Gene Expression in the Susceptibility and Severity of COVID-19. Ann Lab Med 41, 129–138 (2021).

9. Tartof, S. Y. et al. Obesity and mortality among patients diagnosed with COVID-19: Results from an integrated health care organization. Ann. Intern. Med. 173, 773–781 (2020).

10. Marini, J. J. & Gattinoni, L. Management of COVID-19 respiratory distress. JAMA 323, 2329–2330 (2020).

11. Pairo-Castineira, E. et al. Genetic mechanisms of critical illness in COVID-19. Nature 2020 591:7848 591, 92–98 (2020).

12. Patel, D. et al. Machine learning based predictors for COVID-19 disease severity. Scientific Reports 2021 11:1 11, 1–7 (2021).

13. Bai, Y. et al. Presumed Asymptomatic Carrier Transmission of COVID-19. JAMA - Journal of the American Medical Association 323, 1406–1407 (2020).

14. Baillie, J. K. Targeting the host immune response to fight infection. Science (1979) 344, 807–808 (2014).

15. Casanova, J. L. et al. A Global Effort to Define the Human Genetics of Protective Immunity to SARS-CoV-2 Infection. Cell 181, 1194–1199 (2020).

16. Genomewide Association Study of Severe Covid-19 with Respiratory Failure. New England Journal of Medicine 383, 1522–1534 (2020).

17. Lu, R. et al. Genomic characterisation and epidemiology of 2019 novel coronavirus: implications for virus origins and receptor binding. The Lancet 395, 565–574 (2020).

18. Zhang, Q. et al. Life-Threatening COVID-19: Defective Interferons Unleash Excessive Inflammation. Med 1, 14–20 (2020).

19. Onoja, A. et al. An explainable model of host genetic interactions linked to COVID-19 severity. Communications Biology 2022 5:1 5, 1–14 (2022).

20. Onoja, A. et al. An explainable model of host genetic interactions linked to COVID-19 severity. Communications Biology 2022 5:1 5, 1–14 (2022).

21. Miller, T. Explanation in artificial intelligence: Insights from the social sciences. Artif Intell 267, 1– 38 (2019).

22. Ahmad, M. A., Eckert, C. & Teredesai, A. Interpretable Machine Learning in Healthcare. Journal of Machine Learning Research 21, 559–560 (2018).

23. Aggarwal, S., Acharjee, A., Mukherjee, A., Baker, M. S. & Srivastava, S. Role of Multiomics Data to Understand Host-Pathogen Interactions in COVID-19 Pathogenesis. J Proteome Res 20, 1107–1132 (2021).

24. Muhammad, L. J. et al. Supervised Machine Learning Models for Prediction of COVID-19 Infection using Epidemiology Dataset. SN Comput Sci 2, 1–13 (2021).

25. Molnar, C., 2020. Interpretable machine learning. Lulu. com.

26. Molnar, C., Casalicchio, G. & Bischl, B. Interpretable Machine Learning – A Brief History, State-of-the-Art and Challenges. Communications in Computer and Information Science 1323, 417–431 (2020).

27. Kuleshov, M. v. et al. Enrichr: a comprehensive gene set enrichment analysis web server 2016 update. Nucleic Acids Res 44, W90–W97 (2016).

28. Home - Open Targets. https://www.opentargets.org/.

29. Stawiski, E. W. et al. Human ACE2 receptor polymorphisms predict SARS-CoV-2 susceptibility. bioRxiv (2020) doi:10.1101/2020.04.07.024752.

30. Lassau, N. et al. Integrating deep learning CT-scan model, biological and clinical variables to predict severity of COVID-19 patients. Nature Communications 2021 12:1 12, 1–11 (2021).

31. Yan, L. et al. An interpretable mortality prediction model for COVID-19 patients. Nature Machine Intelligence 2020 2:5 2, 283–288 (2020).

32. Doewes, R. I., Nair, R. & Sharma, T. Diagnosis of COVID-19 through blood sample using ensemble genetic algorithms and machine learning classifier. World Journal of Engineering 19, 175–182 (2021).

33. Albadr, M. A. A. et al. Optimised genetic algorithm-extreme learning machine approach for automatic COVID-19 detection. PLoS One 15, e0242899 (2020).

34. Marcos, M. et al. Development of a severity of disease score and classification model by machine learning for hospitalized COVID-19 patients. PLoS One 16, e0240200 (2021).

35. Fallerini, C. et al. Common, low-frequency, rare, and ultra-rare coding variants contribute to COVID-19 severity. Hum Genet 141, 147–173 (2022).

36. Fallerini, C. et al. Common, low-frequency, rare, and ultra-rare coding variants contribute to COVID-19 severity. Hum Genet 141, 147–173 (2022).

37. Sofaer, H. R., Hoeting, J. A. & Jarnevich, C. S. The area under the precision-recall curve as a performance metric for rare binary events. Methods Ecol Evol 10, 565–577 (2019).

38. explainerdashboard · PyPI. https://pypi.org/project/explainerdashboard/.

39. ExplainerDashboard — explainerdashboard 0.2 documentation. https://explainerdashboard.readthedocs.io/en/latest/dashboards.html.

40. oegedijk/explainerdashboard: Quickly build Explainable AI dashboards that show the inner workings of so-called ‘blackbox’ machine learning models. https://github.com/oegedijk/explainerdashboard.

